# Midwifery Practice in Conflict Settings: Lived Experiences from Somalia and Nigeria

**DOI:** 10.64898/2026.06.07.26355130

**Authors:** Emilia Ngozi Iwu, Asia Mohamed Mohamud, Hawa Hawa Abdullahi Mohamed, Charity Maina, Rejoice Helma Abimiku, Sussan Israel-Isah, Kazeem Olalekan Ayodeji, George Odonye, Meighan Mary, Maryan Abdulkadir Ahmed, Salomine Ekambi, Ichchha Pradhan, Hannah Tappis, Shatha Elnakib

**Affiliations:** Institute of Human Virology, Abuja, Nigeria; Rutgers University, School of Nursing, Newark, New Jersey, USA; Somali Research & Development Institute, Mogadishu, Somalia; International Health Department, Johns Hopkins Bloomberg School of Public Health, Baltimore MD USA; Center for Humanitarian Health, Johns Hopkins Bloomberg School of Public Health, Baltimore MD USA

## Abstract

**Background:** Midwives are a central cadre in the health system, particularly in conflict-affected settings where they are sometimes the primary or even only skilled providers available. Yet, despite their critical role, there is limited qualitative evidence capturing their lived experiences and how these shape workforce entry, retention, and overall well-being.

**Methods:** Drawing on a phenomenological research methodology, this qualitative study was embedded within a larger prospective longitudinal cohort of midwifery students and graduates in Somalia and Nigeria. We conducted focus group discussions with graduate midwives (n=48 in Nigeria; n=63 in Somalia) to explore their experiences transitioning into the workforce and their realities working in health systems impacted by conflict and violent insecurity. Data were analysed using inductive thematic analysis.

**Results:** Five themes emerged from the data: (1) job search and workforce entry, which was described as fraught with challenges and shaped by a set of formal systems in Nigeria but informal networks and structural barriers in Somalia (2) working conditions that were marked by resource scarcity, infrastructural challenges, and heavy and unreasonable workloads, (3) safety, security and coping strategies that differed across the two contexts but reflected persistent exposure to violence and a reliance on ad hoc and personal coping in lieu of systematic protection, (4) community perceptions of midwives, shaped and constrained by social and gender norms and (5) mental health and emotional wellbeing, highlighting stress, burnout and moral injury experienced by this cadre.

**Conclusion:** Our findings highlight the profound challenges faced by midwives working in conflict-affected settings, and they shine a light on the urgent need to support and invest in this critical and predominantly female health workforce.

## Introduction

Midwives play a crucial role in improving maternal and newborn health outcomes globally, forming the backbone of care in many health systems [1]. Evidence from a global modelling study suggests that even modest increases in midwife-delivered interventions could avert up to 22% of maternal deaths, 23% of neonatal deaths, and 14% of stillbirths by 2035 [2]. These findings reinforce the importance of strengthening midwifery services, particularly in low- and middle-income countries, where such interventions are most needed [1] [2]. Yet despite this evidence, the global health workforce continues to face a projected shortfall of approximately 310,000 midwives by 2030, a gap most acutely felt in settings already experiencing fragile health systems and high mortality [3].

The urgency of this role is evident in countries like Nigeria and Somalia, which report some of the highest maternal and neonatal mortality burdens globally. Nigeria currently has an estimated maternal mortality ratio of 993 per 100,000 live births, the highest worldwide, while Somalia reports a maternal mortality ratio of approximately 563 per 100,000 live births alongside one of the highest infant and neonatal rates globally [4] [5] [6]. Both countries have regions that have endured prolonged periods of insecurity, resulting in forced displacement, damaged infrastructure, and disrupted health services.

In these contexts, the significance of the midwifery workforce becomes evenmore pronounced because health systems are often weakened, fragmented, or non-functional and midwives are frequently the primary, and sometimes the only, providers of professional maternal and newborn care [7]. These health professionals work under extremely challenging conditions. Health facilities often lack essential equipment, medications, and basic infrastructure, including reliable water, electricity, and adequate space, while persistent shortages of trained personnel force midwives to manage high caseloads with minimal support [8] [9] [10] . These resource constraints contribute to excessive workloads, fatigue, and compromised care delivery. Evidence further suggests that such resource limitations and staffing pressures are associated with increased exposure to workplace violence and occupational hazards, highlighting a concerning link between systemic overwork and provider safety [11] [12].

The psychological toll in these settings extends beyond exhaustion to include “moral distress.” Moral distress is the painful psychological disequilibrium that occurs when providers know the right clinical action to take but are prevented from doing so by institutional constraints or lack of resources [13]. Research indicates that while midwives often possess a deep devotion to their patients, the inability to provide adequate help due to factors beyond their control leads to profound feelings of powerlessness and guilt [14] [15]. Despite the severity of these burdens, the mental health of midwives in crisis-affected regions remains under-researched and under-addressed in global health initiatives.

Several critical gaps persist in the existing literature, including limited qualitative evidence capturing the lived experiences of midwives working in fragile and conflict-affected contexts, particularly in Sub-Saharan Africa. Few studies examine how working conditions, mental and emotional well-being, and career decision-making collectively shape midwife retention and workforce stability. For example, a 2022 review of studies measuring job satisfaction of midwives found most studies focused on workload, working relationships, financial rewards, career development and supervision, but limited and inconsistent attention to midwives’ physical and mental health, or professional autonomy [16]. Another review found only 11 qualitative studies documenting midwives’ experiences in rural areas of Africa, only one of which was conducted in a conflict-affected area (Democratic Republic of Congo) and most of which explored working conditions, emotional distress, social support, and career progression as separate domains rather than interconnected dimensions of workforce stability [8].

This study addresses these gaps by examining the lived experiences of midwives in conflict-affected settings in Somalia and Nigeria, generating context-specific evidence to inform interventions aimed at reducing systemic barriers and strengthening midwife retention in fragile health systems.

## Methods (1699 words)

### Study design

This phenomenological qualitative study [17] was nested within a larger prospective, multi-cohort longitudinal study of midwifery students and recent graduates that commenced enrolment in 2023 and followed participants through 2025. Graduated midwives were drawn from this parent cohort to participate in focus group discussions (FGDs) conducted in October-December 2024 and October-December 2025.

This qualitative study sought to explore and understand the lived experiences of midwives educated and practicing in conflict-affected settings. Phenomenology was selected as the methodological framework to capture the essence and meaning of midwives’ experiences in a context where the intersection of conflict, resource constraints, and professional practice creates unique and complex realities that cannot be adequately understood through quantitative measures alone [18].

The FGDs were designed to elicit rich, in-depth narratives across multiple dimensions of professional experience: training within conflict-affected environments, transition to professional practice, current working conditions, self-assessed clinical competencies, safety and security challenges encountered in service delivery, and evolving professional identity and career aspirations.

### Study Setting

#### Nigeria

The study was conducted in Yobe State in northeastern Nigeria where armed conflict, persistent insecurity, and health system fragility has negatively affected essential health service delivery [19] [20]. Violence, population displacement, and extensive infrastructural damage have led to closure of some health facilities, further impeding access to services especially maternal and newborn healthcare (MNH)[19] [20]. Midwifery training occurs through two primary pathways: 1) a three-year Basic Midwifery programme, which provides comprehensive training for hospital- and community-based maternal and newborn care and 2) the Community Midwifery programme, a 2-year, context-adapted training model approved by Nursing and Midwifery Council of Nigeria in 2018, specifically to strengthen primary healthcare delivery in rural and underserved areas [21] [22]. Both cadres of midwives fulfil critical roles in providing ante prenatal, intrapartum, and postnatal care services. Midwives in Yobe State operate under exceptionally constrained conditions characterized by severe staff shortages, inadequate equipment and supplies, weak referral systems, and elevated security risks [19] [23] [24].

#### Somalia

In Somalia, research was conducted in the Banaadir and Galgaduud regions. Mogadishu is the capital of Somalia and its largest city. For the last three decades, Mogadishu has been struggling with insecurity and violence from conflicts between clan warlords, Al-Shabaab, and the Government [25], while Galgaduud is an administrative region of Galmudug state in the central part of Somalia, which borders Ethiopia. Galgaduud has also faced similar challenges with clan clashes in recent decades, natural disasters, including recurrent droughts, add to the complex humanitarian crisis [26].

Due to the high maternal mortality ratio and severe shortage of health workforce [27], the Ministry of Health (MoH) of the Somali Federal Government and the United Nations Population Fund (UNFPA) established four midwifery training institutions nationwide in 2012 followed by several private universities that provided separate midwifery programs. Since then, UNFPA established 14 midwifery institutions while private universities expanded. midwifery education program is provided through two categories, which are private universities and a community-based program supported by the Ministry of Health and UNFPA. The community-based program includes a direct entry midwifery program lasting three years and a post-nursing midwifery program, which usually takes about one and a half years to complete. These community-based programs enroll midwife students through a targeted recruitment approach, where students are selected through a national enrollment process that ensures that different regions are represented, especially rural and undeserved areas, with the expectation that graduates will return to serve their communities. Meanwhile, the private midwifery programs are offered by private universities; these programs are usually direct entry from secondary school and last three to four years, depending on the university. Private programs are largely urban-based, fee-paying, with less direct linkage to national workforce deployment strategies. This study was carried out with graduates from public and private direct-entry programs, which include a 3-year diploma program and a 4-year bachelor’s program.

### Participants and sampling

In Nigeria, this study was conducted at Shehu Sule College of Nursing and Midwifery, a government-operated institution that educates both basic and community midwives. This institution was purposively selected because it was the first accredited by Nursing and Midwifery Council of Nigeria to implement the newly approved 2-year community midwifery program. The school is located in Damaturu, the capital city of Yobe State.

In Somalia, seven private universities and the only community college in Mogadishu were purposely selected to participate in the cohort study from a list of 16 private universities that offer direct entry to midwifery programs. In Galgaduud, the only midwifery school that provides direct entry was selected.

Study participants were randomly selected from the underlying parent study which followed midwifery students over time. We selected participants who had graduated over two separate rounds of data collection and invited them to participate in focus group discussions (FGDs) to better understand their job-search and work experiences, perspectives on safety and security on the job, and their coping mechanisms in relation to insecurity as well as information about their career plans and perceived ideal jobs.

This longitudinal sampling approach allowed for exploration of experiences at different time points in the graduates’ professional trajectories, capturing both early career transitions and evolving perspectives as participants gained additional practice experience in conflict-affected settings.

### Data collection

In Nigeria, all eligible graduate midwives, who were part of the parent sutdy, were invited through official communication from the Office of the Provost at Shehu Sule College of Nursing Sciences, School of Midwifery . A total of 48 graduates consented and participated in six FGDs conducted in Damaturu (two groups in the first round and four groups in the second round of data collection). Group sizes varied, with seven to nine participants per group. Prior to data collection, trained members of the Nigerian research team, fluent in both English and Hausa, provided comprehensive information about the study, including its purpose, procedures, voluntary nature, confidentiality measures, potential risks and benefits, and the right to withdraw at any time without consequence. Each FGD was conducted by two researchers (CMP-Nurse-midwife/ social worker, and SII-Social scientist and qualitative data analyst) one moderator and one note-taker and lasted between one and two hours.

In Somalia, following each round of follow-up surveys, researchers randomly selected participants who were currently working and contacted them to invite them to participate in FGDs. Those who expressed interest were invited to attend sessions at the Somali Research and Development Institute (SORDI) office in Benadir and atprivate conference hall in Galgaduud. Teams of two trained researchers from SORDI one serving as moderator and the other as note-taker—conducted the interviews in Somali language. They provided participants with detailed information about the FGDs and obtained written informed consent prior to participation. Three FGDs were conducted in Benadir and two in Galgaduud during each round. Across all rounds, a total of sixty three participants took part, with seven participants contributing to two rounds. Sessions lasted approximately one to two hours.

## Data Analysis

All FGDs were audio-recorded with participants’ permission,transcribed and translated by trained transcriptionists. In Nigeria, recordings in Hausa were translated directly from Hausa to English by bilingual transcriptionists with expertise in both languages. In Somalia, recordings in Somali were first transcribed into Somali and subsequently translated into English by certified translators. To ensure data quality, fidelity, and accuracy, trained researchers reviewed all transcripts against the original audio recordings to verify completeness and linguistic equivalence.

Data was analyzed employing inductive coding strategies to capture the essence of participants lived experiences. In Nigeria, three researchers independently coded the transcripts using Dedoose, with coded segments subsequently exported for thematic synthesis. In Somalia, all transcripts were imported into Dedoose qualitative data analysis software (Version 10.0.59, Sociocultural Research Consultants, LLC), where three researchers from Johns Hopkins University and the Somali Research and Development Institute (SORDI) conducted systematic coding.

An initial codebook was developed based on the study objectives encompassing key domains including transition to employment and job search experiences, workplace conditions and organizational environment, safety and security challenges, coping strategies and resilience mechanisms, perceptions of ideal employment, and professional identity and career aspirations. During iterative engagement with the data, the codebook was refined and expanded to incorporate emergent themes grounded in participants’ narratives, including critical shortages of medical supplies and equipment, infrastructural constraints affecting service delivery, and interpersonal challenges involving patients and family members. This iterative process ensured that the analysis remained open to unanticipated dimensions of experience while maintaining focus on the research questions.

### Collaborative Analysis and Triangulation

To enhance analytical rigor and achieve consensus in interpretation, a phased collaborative analysis process was implemented. Initial country-specific in-person workshops were conducted to synthesize data within each context. In Nigeria, researchers from the Institute of Human Virology Nigeria (IHVN) convened an in-person workshop in January 2025 to conduct preliminary thematic synthesis and identify contextually salient patterns. Similarly, an in-person workshop was held in Somalia in March 2025, where two researchers from SORDI and one researcher from Johns Hopkins University engaged in intensive data immersion and initial theme development.

Following these country-specific analyses, a collective cross-country synthesis workshop was conducted virtually in January 2026, bringing together the full multi-country research team: three researchers from SORDI, three from Johns Hopkins University, and six from IHVN. This integrative session facilitated in-depth discussion of coded data, systematic comparison of findings across Nigeria and Somalia, triangulation across datasets, negotiation of divergent interpretations, and consolidation of overarching and context-specific themes. The phased analytical approach moving from country-specific immersion to collective synthesis allowed for both contextual depth and cross-national insight, while the multi-country research team’s diverse expertise enriched interpretation and enhanced the credibility, dependability, and transferability of findings.

### Ethical considerations

Ethical approval for this study was obtained from the Yobe State Ministry of Health Ethics Committee (Reference: No. MOH/GEN/747/VOL.I.), the Somali Research and Development Institute (SORDI-EA02 52) and the Johns Hopkins Bloomberg School of Public Health Institutional Review Board (Reference: No. 00024146).

All participants had previously provided written informed consent during the baseline longitudinal study. Prior to participation in the FGDs, additional verbal consent was obtained, including consent for audio recording. At the beginning of each FGD, an oral consent script was read aloud to reintroduce the study, clarify potential risks and benefits, emphasize voluntary participation, and assure confidentiality. Participants were informed of their right to withdraw at any time without penalty.

## Results

Five overarching themes were identified: (1) job search and workforce entry, (2) working conditions, (3) safety, security and coping strategies, (4) community perceptions of midwives and (5) mental health and emotional wellbeing.

### 1. Job search and workforce entry

Job search, recruitment and deployment experiences differed between the two countries.

In Somalia, many of the graduates described job searching as a largely informal, prolonged, and emotionally taxing process. Personal networks, clan affiliations, and social connections were widely perceived as critical determinants of employment success, particularly in Mogadishu. Contrasting experiences suggests variation in employment pathway; some respondents felt that hiring practices are gradually shifting toward greater emphasis on qualifications and experience. For instance, in Galgaduud, a subset of participants described a more structured, merit-based recruitment process moderated by the regional Ministry of Health. Volunteering was the primary entry route into paid work, with most participants reporting unpaid periods of three to nine months, and in some cases over a year, before securing salaried positions. Graduates from public training institutions were perceived to secure jobs more easily than those from private universities, largely due to longer clinical training, greater exposure to cases, and a willingness to work in rural or underserved areas. In contrast, some participants from private institutions felt disadvantaged because of the shorter practicum periods and limited hands-on experience. Despite Somalia’s acute shortage of trained midwives, employment opportunities appeared sparse; a subset accepted employment in unrelated fields, ranging from infant and young child feeding programs, to translation and insurance work, though most expressed an intention to return to midwifery when circumstances allowed.

> “*Still, there are other issues with job hunting, such as being excluded from the shortlist due to clan [tribal] favoritism or personal bias. At least eight job opportunities that I was professionally qualified for were denied to me, which [saddens me deeply].”* Community Midwife, Galgaduud, *Somalia*
>
> *“Initially, I wanted to work in midwifery, but now I am working in an insurance company. My role involves liaising between hospitals and the company. I prepare reports and facilitate coordination between the two parties.”* Basic midwife, Mogadishu, Somalia

For participants in Yobe State, Nigeria, most graduates entered the workforce through formalized, state-mediated deployment mechanisms; they received official appointment letters before or shortly after graduation and were posted to facilities by the programme director through a standing arrangement with the Yobe State government. This process was more seamless for the community midwives whose program required job appointment as trainees by their local government authorities. The basic midwives whose program did not require trainee appointments expressed frustration with difficulty and lack of control over their posting. Some basic midwives assigned to dangerous areas with security issues experienced further delay as they waited for safer assignments.

> “*We already have job appointments from school so getting a job after school was not a a challenge for us. When we graduate, they post us to our various places of work.”* Community midwife, Nigeria
>
> *“Finding a job is not that hard, we are automatically employed after graduation. The school management and the government have a good relationship in the sense that when an indigene of the state graduates from the midwifery school they get employed.”* Basic midwife, Nigeria

### 2) Working conditions

FGD participants in both countries described chronic shortages of essential medicines and supplies, infrastructure, and human resources that compromised their ability to provide quality care. In Somalia, the stockouts mentioned most often were oxytocin, IV cannulas, sutures, basic lab tests such as haemoglobin measurement, and oxygen machines used to assist patients in respiratory distress. In Nigeria, participants reported shortages of forceps, non-functioning suction machines, and shortages of essential medicines for emergency obstetric and newborn care directly affecting clinical outcomes and causing distress among providers. In both countries, midwives described the most severe supply shortages in rural and public facilities; in Somalia, some midwives reported sending women to private pharmacies and laboratories when supplies were not available, noting concerns that this shifts the cost and burden of responsibility to households.

> *“Yes, it happens a lot. There are times when the facility doesn’t have enough supply, and a mother who is in labour comes to you, and you see she is anaemic, but you don’t have any test to confirm that. So you tell them to go outside and take a blood test. She may refuse, or the family may refuse.”* Community Midwife, Mogadishu Somalia
>
> *“Sometimes, we deliver a baby with severe asphyxia and secretion, and we want to clear the airway, but there is no suction machine. It is there, but it’s not functioning.”* Basic midwife, Nigeria

Infrastructure challenges compound stresses of supply shortages. Midwives in both countries described having to conduct deliveries by torchlight or mobile phone screen due to unreliable electricity. In Somalia, some midwives also described working in facilities that had one delivery set, which had to be transported to the town for sterilization, and working in sites where ambulances were unavailable or delayed, causing life-threatening situations in obstetric emergencies. Similar conditions were reported in Nigeria where participants described poor facility infrastructure, as well as unsafe and poorly maintained staff accommodations without protective fencing.

Understaffing also amplified the challenges of providing services with limited resources. Midwives in both countries described managing maternity wards or entire facilities alone, sometimes managing multiple critically ill patients simultaneously. In Nigeria, some participants also expressed deep frustration with low salaries, lack of allowances, and limited access to training opportunities.

> “*I am the only midwife working there. We don’t have enough manpower and there is no nearby facility so people in the community are all coming there so the workload becomes too much for us.”* Community midwife, Nigeria

Socio-cultural norms and dynamics of interactions with maternity clients and their families added another layer of complexity to working conditions. In Somalia, young midwifery graduates noted policies and cultural norms granting male family members decision-making authority led to refusals of medically indicated interventions, including caesarean sections, blood transfusions, and episiotomy repair, sometimes with fatal consequences. FGD participants reported frequent interference from the relatives during care, including refusal to leave delivery rooms, mistrust of clinical procedures, accusations of incompetence due to their age, which resulted in requests for “older” or “senior” midwives, and verbal and physical abuse. In Nigeria, cultural norms and beliefs were reported as major barriers to providing timely and appropriate care. Midwives described cases in which women with severe postpartum haemorrhage and anaemia refused food, fluids, and medication due to traditional beliefs that eating or drinking before performing specific cultural rituals could harm or kill the newborn. In both settings, chronic understaffing and heavy workloads limited midwives’ capacity to invest the time needed to explain clinical reasoning, negotiate care options with families, and de-escalate conflicts; as a result, refusals of care sometimes led to preventable adverse outcomes.

> *“A postoperative patient who was nil per os (not allowed to take anything by mouth) was brought in, but her family gave her water. The team I work with went to them and explained that the patient was not allowed to drink water and asked them to stop; otherwise, her condition may become serious. The male relatives accompanying the patient became angry, brought a drip-stand, and hit the female staff with it.”* Basic Midwife, Mogadishu, Somalia
>
> *“For instance, there are cases where a mother is in labor, and the baby has run out of amniotic fluid, necessitating a caesarean section or induction. However, the family members refused these interventions, and they returned to their home. After three days, they came back with intrauterine fetal death (IUFD). This situation arises from the family’s decision-making authority.”* Community Midwife, Galgaduud, Somalia
>
> *“The woman had postpartum haemorrhage and Packed Cell Volume of 20%, but she refused to eat, drink, or take medication because they believed if she eats the baby will die.”* Basic midwife, Nigeria

### 3) Safety, security and coping strategies

Safety and security concerns shaped many aspects of midwives’ working experiences. In both countries, participants associated the presence of on-site security guards with safety within facility compounds and expressed feelings of insecurity and fear of thieves or youth gangs when working and commuting at night. Beyond these commonalities, descriptions of safety and security threats distinctly differed by location. In Somalia, urban and semi-urban facilities were perceived as relatively safe in comparison to facilities in rural areas where Al-Shabab activity and inter-clan clashes were noted risks. A small number of participants also expressed fear of men coming into the health facility under false pretense of requesting assistance with a woman in labour outside of the facility and then luring the midwife into an unsafe situation. In contrast, while some midwives in Nigeria reported feeling safe at facilities, most did not, largely due to a lack of security fences, gates and guards on site. Several participants described armed raids, kidnappings, Boko Haram incursions, and community violence forcing them to suspend night duties, restrict movement, or rely on volunteers for protection.

> *“The place where I work has good security. Sometimes there are a few robbery cases that happen at night. There are men on motorbikes that snatch your phone, but it happens during late at night.”* Basic Midwife, Mogadishu, Somalia
>
> *“I work at [Facility X]. It faces insecurity and the facility was attacked before and now we don’t have a gate, and the staff quarter is not safe for me to stay. Sometimes, I go into the community to sleep in one of the community leaders’ houses. The facility is no longer fenced.”* Community midwife, Nigeria
>
> *“*…*So, for the past month, we have been at home because Boko Haram came and stole the ambulance and packed some drugs where we were and the place has become very unsafe.”* Basic midwife, Nigeria

Midwives in both countries described both pre-emptive and reactive coping strategies. In Somalia most participants spoke about growing up in an environment of ongoing explosions, chaos, clashes, and insecurity, over time, and adapting to these conditions. Most participants stated that their workplaces are safe, and that they had not personally encountered security issues. Other participants, particularly those who experienced attacks, mentioned that they fled health facilities before or during the attack; others with fear of street violence stated that they refused to leave the health facility at night unless they are escorted by a security guard. In Nigeria, where fear of attacks was more pervasive and facilities perceived as direct targets, midwives reported requesting personal escorts, sleeping within the community during periods of heightened risk, avoiding night shifts entirely, and wearing civilian clothing to avoid identification by armed groups.

> *“Yes, when I was working in the village, there were nights when the internet and communication were cut off. People were fleeing from the village, and I felt very scared because we had to cross the river to reach the safe part of the village, and I did not know how to swim.”* Community Midwife, Galgaduud, Somalia
>
> *“The reason is that we were born and grew up here, so we already adapted to the insecurity itself.”* Community Midwife, Galgaduud, Somalia
>
> *“ so, for me I’m not from this town so the way I cope is anytime I don’t feel safe and I get a call I normally tell them I don’t feel safe so they should send someone who will come to my house and accompany me to the facility when am done the person accompanies me back to my home.”* Community midwife, Nigeria

### 4) Community Perceptions of Midwives

In Somalia, most participants stated that young midwives often face scepticism from care seekers due to their age and perceived inexperience; community members associate maternal experience with midwifery skills, believing that midwives cannot give advice if they do not have children themselves. Critically, this skepticism can be overcome; participants noted that when women experienced respective care and positive birth outcomes, they were grateful, expressed satisfaction, and returned for care in future pregnancies. A generational shift was also noted, with younger mothers seeming more comfortable being attended by midwives closer to their own age.

> *“People don’t trust us until they have seen our work; we even have repeat patients who, at first, didn’t believe us.”* Community Midwife, Mogadishu, Somalia
>
> *“I have a lot of mothers who come looking for me when they want to deliver now they have confidence in us because they have seen me, and also a lot of mothers giving birth are young people; unless their parents force them to be delivered by TBAs, they don’t prefer it, and they come to the MCH regularly.”* Basic midwife, Nigeria

In Nigeria, young midwivest reported facing skepticism because of their age. They explained that communities’ perceptions of midwives were closely tied to their age and qualifications. Participants noted that graduates of the three-year Basic Midwifery programme are widely respected, trusted, and valued, sometimes to the point of creating unrealistically high expectations. Graduates of the two-year Community Midwifery programme, by contrast, were less respected, stating that other health workers looked down on them and frequently regarded them as inferior to nurses. Unlike in Somalia, these perceptions were not overcome when quality care was provided; in fact, some midwives reported that referrals to other facilities were frequently misunderstood and perceived as a reflection of the midwife’s incompetence or unwillingness to work. For a subset of participants, the lack of respect and recognition from community members led them to consider leaving the profession or pursuing further education in nursing or medicine to gain legitimacy and respect within both the hospital and the community.

> *“*.. *In the community they praise midwives so well that they assume you know everything even if you’re focused on delivery, they assume you know also where we lack experience, we refer them to the appropriate person.”* Basic midwife, Nigeria
>
> *“The way they [i.e. other workers] look down on us because we are community midwives is bad because it affects us making us want to leave the job and the truth is we work harder than the basic midwives and the nurses they say we are useless.”* Community midwife, Nigeria
>
> *“I will change the profession because of how the community and senior health worker perceive midwives, they see us as second class workers like we don’t have value at the workplace.So, I want to go back to school to become a nurse or doctor so I can be respected in the hospital and in the community.”* Community midwife, Nigeria

### 5) Mental health and emotional wellbeing

Midwives in both countries described ways in which cumulative stress of job searches, working in understaffed, under-resourced and insecure settings, tensions between clinical duties and family responsibilities, and limited institutional support took tolls on their mental health and well-being. For example, in Somalia, a few participants acknowledged that prolonged unemployment periods after graduation negatively affected their emotional well-being which led to stress, frustration, depression, and feelings of hopelessness. In Nigeria, several midwives reported a state of constant hypervigilance necessitated by the threat of regional insurgency, where the act of commuting involved navigating active conflict zones and fear of kidnapping. In addition to sadness and hypervigilance, common examples of strains on well-being described by midwives in both countries included sleep deprivation, appetite changes, emotional instability, increased irritability and withdrawal from social and family relationships.

> *“I used to be someone who stayed connected with relatives, but now I do not keep in touch with anyone because I feel I do not even have time for myself. Sometimes, I feel psychological pressure and wonder whether I am becoming a bad or irritable person.”* Community Midwife, Mogadishu, Somalia
>
> *“I avoid going outside and stay inside because I feel afraid even when I am hungry; I continue working; the night shift affected my appetite and caused sleep problems. I have also noticed increased forgetfulness during my menstrual period; I experience pain, tiredness, and become inactive and lethargic.”* Basic Midwife,Mogadishu Somalia

In Somalia, witnessing maternal and neonatal loss among women with severe complications, and managing emergencies with inadequate support left many participants feeling sad, helpless, and emotionally exhausted, particularly when patients refused referrals or lacked the financial means to access care. Exposure to traumatic clinical events, including severe haemorrhaging and birth injuries, produced lasting trauma that manifested as inability to eat or sleep. Many participants in both countries described a process of desensitization or “adaptation” over time. A more lingering form of distress identified was moral injury, where the midwives felt “haunted” by mistakes made due to exhaustion.

> *“Sometimes you intend to do the right thing but due to exhaustion you end up not doing it right. You feel guilty knowing you did something wrong*… *it keeps haunting you until gradually you forget it.”* Basic midwife, Nigeria
>
> These burdens extended beyond the clinical setting into midwives’ personal and family lives. In Nigeria, midwives with spouses and children struggled with guilt when professional obligations took them away from family responsibilities, while some unmarried participants described anticipatory anxiety about future family formation and work-life balance.
>
> *“I’m always tired because I am the only one. Be it morning, afternoon, or night shift it must be me*… *I get tired sometimes. I spend four to six weeks at my post; my husband must be the one to come see me. I don’t get time to rest.”* Community midwife, Nigeria
>
> “*For me the fear is what if I get married how will l cope with the stress of work coming back and handing the shifts everyday … I don’t know how I’ll cope with that”* Basic midwife, Nigeria
>
> *“By the time I get to the school, the break time is already over with my children looking at others eating like beggars; it makes me sad that I had to do that to them*… *this profession is not for someone with children.”* Basic midwife, Nigeria

Despite these pervasive challenges, midwives in both countries identified sources of encouragement and motivation. Some participants mentioned that positive patient outcomes, supportive colleagues, and professional recognition —including encouragement from senior midwives and obstetricians in Somalia — enhanced their sense of fulfilment, confidence, and emotional resilience.

## Discussion

Our study sheds light on the lived realities of midwives working in conflict-affected settings in Somalia and Nigeria, highlighting the convergence of insecurity, fragile health systems, and gendered expectations in shaping their professional and personal lives. Taken together, our findings point the important role that midwives play in maintaining health systems under strain, but they also reveal the significant burden and precarity experienced by this predominantly female workforce.

In Nigeria, midwives described relatively smoother entry into the workforce, specifically for community midwives where a formalized, state-sponsored recruitment and deployment process facilitates their transition from school to employment. In Yobe State, coordination between training institutions and government authorities enabled the absorption of new graduates into the public sector, reducing uncertainty for community midwives.

Although such regulatory approaches including pre-arranged deployment and return-of-service requirements are often viewed as effective solutions to workforce shortages in rural and underserved areas, evidence on their effectiveness is still thin and mixed [28]. In some contexts, these interventions have been associated with low rural retention once mandatory service periods end, while in others, deployment to source communities has improved workforce availability. In Afghanistan, for example, Zainullah et al. (2014) found that pre-arranged deployment increased employment outcomes for midwives [29]. In Nigeria, many participants in community midwifery programs hail from the same remote and conflict-affected areas experiencing shortages, which according to existing evidence can be associated with improved rural retention across health professions [12] [30] [31] [32] [33]. However, the extent to which these strategies are effective for midwives in conflict-affected settings, and whether they work on the long run in retaining midwives, still remains underexamined.

In contrast, in Somalia, midwives, particularly those working in the private sector, relied heavily on personal networks and clan affiliations to secure work, despite a well-documented national shortage of midwives [23]. Participants described their job searches as prolonged and uncertain, often involving extended periods of unpaid volunteering and reliance on social connections and familial ties. While some participants spoke about recent shifts toward more merit-based recruitment, employment pathways remained precarious and were described as emotionally taxing and demoralizing. These experiences echo broader evidence from LMICs which shows that workforce entry is not only shaped by training pipeline and ‘need,’ but by other factors that can delay or prevent absorption of newly trained health workers even in contexts characterized by health worker shortages [34]. Our findings make the case that recruiting from underserved areas and linking training to local deployment can be a critical measure to improve retention, but this must be accompanied by transparent, merit-based recruitment systems and stronger transition-to-work pipelines, particularly in contexts like Somalia where this was identified as a key gap.

Consistent with evidence from other conflict-affected settings, safety and security challenges were salient themes emerging in our data, and midwives experienced the insecurity not only in their personal but also professional lives [13][29] . While experiences were similar in many ways across the two countries, with similar coping strategies used, there were distinct differences in how violence manifested, which necessitate different approaches and strategies for the protection of health workers. In Nigeria, midwives reported acute exposure to insurgent violence, including facility attacks, kidnappings, and armed ambushes, particularly in rural postings. The absence of physical security infrastructure -- such as fences, gates, guards, or safe staff accommodation -- left many feeling unprotected, especially during night shifts. Somali midwives, by contrast, generally perceived urban facilities as relatively safe, but emphasized risks associated with commuting, night travel, and rural deployment, including Al-Shabaab activity, clan clashes, and gang violence. In Somalia, protective measures that extend beyond the facilities are needed; these can include measures that facilitate access to safe transport as well as engagement of community members in protection of health staff. In Nigeria, basic infrastructure (e.g., lighting, fencing) are more urgent investments. Together, these findings underscore the need for interventions that may fall outside of traditional humanitarian health responses, such as safe transport and community-based protection mechanisms, but that are critical for ensuring both safety and continuity of care.

Across both countries, participants relayed context-specific coping strategies but those were largely ad hoc and reactive, rather than systematic or institutional. They included avoiding night shifts, relying on community escorts, avoiding uniforms to prevent identification by militant groups, embedding oneself within communities. The pervasiveness of insecurity in many areas where the participants worked and their reported reliance on personal coping mechanismsunderscore the urgent need for systematic institutional protections for these very vulnerable workers. While locally-sourced coping strategies, like the ones we describe in our study, may be effective and applicable in other contexts, they must be coupled with institutional and systematic protections, at the local, national and global level to ensure that health workers are protected in conflict settings. Moreover, the psychological toll of these conditions was profound and consistent across the two settings. The impact that insecurity, excessive workload, and shortage of resources has on the mental health and wellbeing of these midwives echoes broader literature on the experiences of frontline workers in conflict settings. Witter et al document high levels of exhaustion, psychological strain and burnout among health workers in conflict [35]. Dean et al find that during moments of resource constraints, healthcare workers in Liberia and Sierra Leone spoke about the guilt and shame they felt for making decisions due to scarcity that challenged their morals and values, mirroring the moral injury felt by midwives in our study who reported an inability to provide adequate care not due to lack of training or interest in patients’ wellbeing but rather due to the shortage of resources and structural constraints [36]. This distress is further intensified by gendered role expectations: our study aligns with studies in other settings that show that female health workers -- especially married women and mothers -- experience compounded pressure as they attempt to reconcile demanding work with social expectations tied to their gender [37]. The psychological burden experienced by midwives echoes broader evidence of burnout and trauma in conflict settings; alongside improving working conditions, low-cost interventions such as peer and group-based psychosocial support can help ameliorate the conditions under which midwives operate[38], [39].

This study is not without limitations. First, our analysis draws on the experiences of midwives who have either entered the workforce or continue to seek employment as midwives. We did not include those who have exited the profession, whose perspectives may be profoundly different and who could offer additional insights into attrition. Second, our sample is drawn from specific locations in Nigeria and Somalia, and caution should be exercised in generalizing findings across all humanitarian settings, as many of the challenges identified are context specific. Nevertheless, we observed important commonalities across the two settings, especially in relation to the demanding and often precarious working conditions faced by midwives, which suggest broader relevance to similar conflict-affected contexts. Finally, although we employed local data collectors familiar with the context, it is possible that participants self-censored or refrained from sharing sensitive experiences. Recall and social desirability bias may have also influenced participants’ accounts.

Taken together, our findings paint a textured picture of the lived realities of midwives operating in conflict settings, where health workforce shortages are acute and this cadre is critically needed, yet where structural and contextual challenges undermine their entry into, motivation, and retention in the health workforce

## Conclusion

This study contributes qualitative evidence on a workforce that is simultaneously indispensable and deeply under-protected in some of the world’s most challenging health system contexts. The lived experiences documented here reveal midwives holding fragile systems together under conditions of violence, resource scarcity, and institutional neglect, often without formal recognition or support. Across Nigeria and Somalia, common systemic drivers emerge with striking consistency — structural under-investment in security and infrastructure, inadequate human resource support, and the absence of psychosocial safety nets for a predominantly female workforce operating at the intersection of professional obligation and gendered social expectation. Strengthening midwifery workforces in these environments demands a reorientation of policy attention: from exclusively focusing on how many midwives can be trained, toward whether the conditions in which they work are viable and sustainable. Transparent and merit-based recruitment, community-embedded security mechanisms, and structured psychosocial support are foundational investments in a functional midwifery workforce. In settings where a midwife may be the only skilled professional a woman encounters across her entire reproductive life, her sustained presence in the system is a matter of survival.

## Data Availability

All data underlying this analysis is available on reasonable request from the corresponding author

## Declarations

## Ethics approval and consent to participate

Ethical approval was obtained by John Hopkins Bloomberg School of Public Health (IRB00024146) and the Somali Research and Development Institute (SORDI-EA02 52) and Yobe State Health Ministry of Health Ethics Committee (MOH/GEN/747/VOL.I) All procedures, including obtaining written informed consent from participants, will strictly comply with applicable regulations and the Helsinki Declaration of 1975.

## Consent for publication

Not applicable

## Author contributions

HT, EI – funding acquisition; HT, EI, HA, CM, AM, SE – conceptualization; HT, EI, HA, CM, AM, SE – methodology; EI, HA, SE – project administration; EI, AM, HA, CM, RA, SI, KA, GO, MM, MA, SE, IP – (data analysis); EI, SE and AM – writing (first draft); HT, EI, HA, CM, AM, SE – writing (review and revision).

## Acknowledgements

The authors acknowledge the midwifery students and graduates who volunteered their time to be part of this study. We also acknowledge the valuable time and effort made by the EQUAL Research Programme Consortium members at the Institute for Human Virology, Nigeria (IHVN), Somalia Research and Development Institute (SORDI), Johns Hopkins Center for Humanitarian Health, as well as to Alicia Adler at the International Rescue Committee for graphic design support.

## Availability of data and materials

All data underlying this analysis is available on reasonable request from the corresponding author

